# Understanding Public Attitudes Towards Human Papillomavirus Vaccination in Japan: Insights from Social Media Stance Analysis Using Large Language Models

**DOI:** 10.1101/2024.10.07.24315018

**Authors:** Junyu Liu, Qian Niu, Momoko Nagai-Tanima, Tomoki Aoyama

## Abstract

**Background:** Despite the reinstatement of proactive human papillomavirus (HPV) vaccine recommendations in 2022, Japan continues to face persistently low HPV vaccination rates, posing significant public health challenges. Misinformation, complacency, and accessibility issues have been identified as key factors undermining vaccine uptake.

**Objective:** This study aims to understand how factors such as misinformation, public health events, and attitudes toward other vaccines, like COVID-19, influence HPV vaccine hesitancy, by analyzing the evolution of public attitudes towards HPV vaccination in Japan by examining social media content.

**Methods:** We collected tweets related to HPV vaccine from 2011 to 2021. Traditional natural language processing (NLP) methods and large language models (LLMs) was utilized to perform stance analysis on collected data. The analysis included stance identification, time series analysis, topic modeling, and logic analysis. We framed our findings within the context of the WHO’s 3Cs model—Confidence, Complacency, and Convenience.

**Results:** Public confidence in the HPV vaccine fluctuated in response to government policies and media events, with misinformation playing a critical role in eroding trust. Complacency increased following the suspension of recommendations in 2013 but decreased as advocacy resumed in 2020. Accessibility (Convenience) was also found to be a key determinant of vaccination uptake. HPV vaccines are often used as supportive evidence towards other vaccines, such as COVID-19.

**Conclusions:** Our findings underscore the importance of targeted public health interventions to restore and maintain vaccine confidence in Japan. While vaccine confidence has shown a slow increase, sustained efforts are necessary to secure long-term improvements. Confidence in one vaccine may positively influence perceptions of other vaccines. Addressing misinformation, reducing complacency, and enhancing vaccine accessibility are key strategies to improve uptake. Increased confidence in HPV vaccines appeared to have a positive influence on confidence in other vaccines, such as COVID-19. This study also demonstrates the utility of LLMs in offering a deeper understanding of public health attitudes. To effectively combat vaccine hesitancy and improve coverage, interventions must prioritize consistent communication, localized strategies, and an integrated approach to vaccine narratives.

## Introduction

In 2020, the World Health Organization (WHO) launched a strategy to eliminate cervical cancer, aiming for all countries to achieve a 90% coverage rate for the human papillomavirus (HPV) vaccination by 2030[1]. In 2023, WHO released estimates of the first-dose vaccination coverage for females in 133 countries, with a global average of 62%, and 15 countries have already met WHO’s target [2]. Amid the context of HPV vaccination in high-income countries, Japan is in a critical situation. In Japan, the bivalent HPV vaccine was licensed in October 2009, and the quadrivalent vaccine in July 2011. Public subsidies for girls in grades 7 to 11 began in 2010 [3]. Starting in April 2013, both bivalent and quadrivalent HPV vaccines were included in Japan’s national immunization program for girls aged 12-16 [4]. However, after widespread reports of adverse events related to HPV vaccination, the Japanese Ministry of Health, Labour and Welfare (MHLW) announced the suspension of its proactive recommendation for the HPV vaccine in June of the same year [5,6], leading to an immediate and sharp decline in public trust and acceptance of the HPV vaccine [7–11] and significantly increased cervical cancer incidence [12,13]. Larson et al. reported that the longer the recommendation suspension in Japan lasted, the greater the public’s concern grew [11]. The WHO Global Advisory Committee on Vaccine Safety commented on the situation in Japan in 2015, stating, “policy decisions based on weak evidence, leading to lack of use of safe and effective vaccines, can result in real harm.” [14]

Recent studies reveal that vaccine hesitancy in Japan has seen only marginal decrease since the proactive HPV vaccine recommendations reinstated in 2022 [15–19]. Despite government efforts, the vaccination rate remains significantly below pre-2013 levels [19]. Several factors have contributed to this slow recovery. Tarada et al. and Lelliott et al. suggest that misinformation and persistent public fears regarding vaccine safety remain key obstacles [16,20]. Ueda et al. reported negative media coverage from the 2013 suspension continuing to affect perceptions [21]. However, localized initiatives, such as those implemented in Shiki City [18], have demonstrated that regional government support, public funding, and targeted campaigns can lead to substantial increases in vaccine uptake. Miyagi et al. emphasize the role of transparent communication and consistent public health messaging, which are crucial in mitigating the effects of past misinformation [17]. These factors all relate to the influence of multimedia, particularly social media, in recent years.

Social media plays a significant role in shaping public perceptions of the HPV vaccine, and recent studies have highlighted both its positive and negative impacts. For instance, Dunn et al. found that exposure to negative information about the HPV vaccine on social media was associated with lower vaccine coverage, underscoring the influence of media controversies on public acceptance [22]. Similarly, Teoh et al. emphasized that social media platforms can both effectively communicate HPV vaccination recommendations and serve as conduits for misinformation, affecting public attitudes and behaviors [23]. In contrast, a study by Pedersen et al. demonstrated that strategic social media campaigns focusing on positive messaging significantly improved engagement rates and public support for HPV vaccination in Denmark [24]. These studies collectively indicate that social has the potential to either promote vaccine uptake through well-targeted campaigns or amplify vaccine hesitancy through the spread of misinformation. This dual nature of social media’s influence is particularly relevant in Japan, where negative media coverage has had a profound impact on public attitudes towards HPV vaccination [21,25,26]. To help release the vaccine hesitation, understanding the public attitude towards HPV vaccine though social media analysis is an import topic in Japan.

Recently, NLP methods are applied to the social media analysis, making social media analysis more accurate and can catch nuanced information from large amount of data [27]. Beside the traditional NLP methods like n-gram [28] and topic modeling [29], Deep learning (DL) models are used for classification tasks, especial sentiment analysis in social media [30–33]. Tomaszewski et al. built a DL model to surveillant misinformation in social media about HPV [34]. As the LLMs become main stream of DL, there are also works starting to analyze social media with LLMs [35].

In this study, we aim to understand the persistently low HPV vaccination rate by analyzing the evolution of public attitudes towards HPV vaccination in Japan between 2011 and 2021. We utilize NLP models to evaluate social media content before the proactive recommendation was reinstated. Our analysis examines the dynamics of vaccine confidence over time, the relationship between public health events and shifts in public opinion, and the interplay between attitudes towards HPV and COVID-19 vaccines. By combining stance analysis, time series analysis, topic modeling, and logic analysis, we seek to provide a deeper understanding of vaccine confidence and the factors that drive vaccine uptake.

## Methods

### Data Collection and Preprocessing

This study utilized the Twitter (now X) API to collect Japanese tweets related to the HPV vaccine between 2011 and 2021. Tweets were retrieved using various Japanese keywords for the HPV vaccine. The number of tweets generally increased each year, resulting in a total of 228,376 tweets. After excluding tweets with unrecognizable dates, 228,300 tweets were retained for subsequent analysis. Following data collection, the tweets underwent cleaning and preprocessing. Retweets were removed using the Python package tweepy [36]. Web links, special characters, emojis, and ampersands were also eliminated, and full-width English characters were converted to half-width, lowercase characters.

### Annotation

To facilitate model training and fine-tuning, 2.5% of the total tweets per year were randomly selected for annotation. This study focused on public stances towards the HPV vaccine, employing a three-category annotation system: advocate, oppose, and unknown. Detailed category definitions are provided in Table 1. The annotation team consisted of four medical professionals, three of whom independently annotated each tweet. Tweets with unanimous annotations were classified as Tier 1, those with two matching annotations as Tier 2, and those without any matching annotations as Tier 3. Tier 1 data, deemed to have the highest consistency, utilized the initial annotations as the final labels. Tier 2 and Tier 3 data underwent further consensus meetings, with final labels determined through arbitration. A total of 5,758 tweets were annotated, with 5,050 in Tier 1, 701 in Tier 2, and 7 in Tier 3. The distribution across the advocate, oppose, and unknown categories in the annotated data was 1,646, 697, and 3,415, respectively.

**Table 1.** Definitions of stances to the HPV Vaccine.

| Category | Definition | Mock tweet example |
| --- | --- | --- |
| advocate | Affirmatively advocacy of the HPV vaccine, highlighting its efficacy in preventing HPV-related diseases, the critical role of vaccination in public health, and positive vaccination outcomes. These messages distinctly advocate for the HPV vaccine's utilization, reinforcing its importance in health prevention measures. | Just got my HPV vaccine today! Feeling grateful for the science that protects us from cervical cancer. Highly recommend it to everyone eligible! |
| opposite | Explicitly challenge or critique the Human Papillomavirus (HPV) vaccine, focusing on concerns related to its efficacy, safety, or potential adverse effects. The critical perspective must be distinctly directed at the HPV vaccine in isolation, excluding broader criticisms of vaccines at large, political deliberations concerning vaccination policies, or financial considerations associated with vaccine procurement. | I'm skeptical about the HPV vaccine due to the side effects I've read about. More research is needed before making it a widespread recommendation. Let's be cautious. |
| unknown | Neither explicitly endorse nor directly oppose the HPV vaccine. This category encapsulates tweets that voice opposition stemming from broad anti-vaxxers, policies, or concerns about the cost, without specifically expressing the advocate or opposite of the HPV vaccine itself. It also includes tweets that are too ambiguous, neutral, or irrelevant to ascertain a clear position concerning the HPV vaccine. | Why isn't the HPV vaccine free for everyone? If it's so important, shouldn't the government cover the cost? Still trying to understand the policy behind this. |

### Stance analysis with deep learning models

#### Model Selection

To achieve optimal results, we first selected the best-performing model. We compared several traditional NLP classification models and LLMs. Among traditional models, we considered long short-term memory (LSTM) [37], Bidirectional Encoder Representations from Transformers (BERT) [38], and DistilBERT [39]. For LLMs, we tried on Gemma-2-2b [40], Gemma-2-9b, and Llama-3.1-8b [41]. We also evaluated Gemini Pro 1.0 [42] from Google AI Studio during free-trial.

For the LSTM model, we designed a two-layer bidirectional architecture. Tweets were tokenized using the Python package Ginza [43]. Due to the manageable parameter size of this model, we trained it from scratch. For the BERT models, we selected Tohoku BERT [44] and Line DistilBERT [45] as baselines. A classification head was added to the first token of the final hidden layer of these models, and fine-tuning was performed using our tweet data, while keeping the BERT parameters frozen.

For the LLMs, computational resource constraints prevented us from fine-tuning models with more than 10 billion parameters, and budgetary constraints precluded fine-tuning using APIs such as ChatGPT [46] or Claude [47]. Therefore, we opted for smaller open-source LLMs and tested both baseline and Quantization and Low-Rank Adaptation (QLoRA)-fine-tuned versions on the tweet data [48]. For Gemini Pro 1.0, we directly finetuned the model within the Google AI studio. For all LLMs, we used the following prompt in Japanese, followed by the original tweet as user prompts:” Does this tweet express a position against the HPV vaccine itself? Please classify this tweet as ‘advocate’, ‘oppose’, or ‘unknown’. Do not make subjective assumptions; base your decision solely on the content of the tweet. Answer only your position, no explanation is required.”

Given the significant class imbalance in the dataset, the 5,758 annotated tweets were split into training, validation, and test sets, with the proportion of each category maintained across the splits. To ensure a robust evaluation, 100 tweets from each category were randomly selected to form the test set. The remaining data within each category was then partitioned into training and validation sets at a ratio of 4:1. Due to the training sample size limitation (<500) imposed by Gemini 1.0 Pro, a subset of 498 labeled tweets (distributed equally across the three categories at a ratio of 166:166:166) was utilized for fine-tuning the model for the HPV vaccine stance classification task. Model performance was assessed using standard evaluation metrics, namely precision, recall, and F1-score. Details about the labeled dataset are shown in Table 2.

**Table 2.** number of labeled tweets of different stances in the dataset.

| category | train set | validation set | test set |
| --- | --- | --- | --- |
| advocate | 2652 | 663 | 100 |
| oppose | 477 | 120 | 100 |
| unknown | 1236 | 310 | 100 |

To further investigate the performance boundaries of the optimal model, we explored the impact of varying category distributions within the training data. We experimented with different ratios of advocate, oppose, and unknown categories, specifically 166:166:166, 150:200:150, 125:250:125, 100:300:100, 75:350:75, and 50:400:50. For each ratio, data was randomly sampled three times, and the model’s performance was evaluated, with the results averaged to ensure robustness.

Upon identifying the optimal category ratio, we fine-tuned the best-performing model five times using this ratio, each time with a new random data sample. The remaining labeled data was utilized for performance evaluation, and the three models exhibiting the highest performance were selected for final inference. For each unlabeled tweet, these three models generated independent predictions. If two or more models agreed on the predicted label, that label was assigned to the tweet. In cases where all three models produced different predictions, the tweet was classified as “unknown.”

#### Time series analysis of stances

Following the stance classification of all tweets, a comprehensive time series analysis was performed on the extracted data to examine the evolution of public sentiment towards the HPV vaccine in Japan between 2011 and 2021. The difference between the monthly counts of tweets advocating for and opposing the HPV vaccine was then calculated to capture the dynamic shifts and relative strength of each stance over time.

To pinpoint significant structural breaks within the time series data, the Pruned Exact Linear Time (PELT) algorithm [49], a computationally efficient method for change point detection, was employed. These identified change points corresponded to major shifts in public opinion regarding the HPV vaccine, potentially linked to specific events or information dissemination campaigns. By juxtaposing the time series analysis results with a timeline of key news events and public health policies, a comprehensive interpretation of the observed trends was achieved. This approach facilitated a deeper understanding of the factors influencing public acceptance of the HPV vaccine and provided empirical evidence to inform future public health interventions and policy decisions.

### LDA topic modeling analysis

In this study, we employed Latent Dirichlet Allocation (LDA) [49] topic modeling, a widely used technique for extracting latent thematic information from large volumes of text data, to analyze the Japanese tweets related to the HPV vaccine between 2011 and 2021. We aimed to gain a deeper understanding of the specific perspectives held by Japanese Twitter users across different stances towards the HPV vaccine by LDA modeling. To determine the optimal number of topics that best represented the thematic structure of the dataset, we experimented with various topic numbers ranging from 1 to 50. For each configuration, we evaluated the model’s performance using perplexity and coherence scores [50]. Details about optimal topic number selection are in Figure 1, 4, 7 of appendix 4. After selecting the optimal number of topics, we conducted further qualitative analysis to refine and interpret the final thematic content. Similar to Niu et al. [51], we calculated the monthly average expectation of tweets belonging to different topics for fine-grain time series analysis.

**Error! Figure 1.** Weekly number of tweets with “COVID-19” keywords, using HPV vaccine as example to share stance to COVID-19 vaccine (“HPV to COVID”), using COVID-19 vaccine as example to share stance to HPV vaccine (“COVID to HPV”), or neither (“Not related”). Left: result on tweets with advocate stance; Right: result on tweets with opposed stance.

In line with prior research indicating that misinformation is an important factor for vaccine hesitation [20,23,52], we also investigated the impact of misinformation on the HPV vaccine. First, we identified topics that were likely to contain misinformation through group discussion, focusing on tweets with an estimated probability of over 80% of belonging to those topics. We then utilized the Claude-3-opus model [47] to assess whether each tweet was attempting to disseminate misinformation regarding the HPV vaccine. To validate the accuracy of the model’s classification, we randomly selected 25 tweets categorized as misinformation and 25 categorized as non-misinformation by the model. Three independent volunteers evaluated the credibility of the information in these tweets by cross-referencing reputable news sources and data from the MHLW [53]. The verification process was repeated three times to estimate the model’s classification accuracy.

### Explore relationship between HPV vaccination and COVID-19

Considering the chronological overlap between the duration of data collection and the COVID-19 pandemic, a significant historical health event, this research further investigated the potential relationship between HPV vaccination and COVID-19. Specifically, the objective was to determine whether the COVID-19 pandemic had an impact on public attitudes towards HPV vaccination. From our dataset, all tweets comprising the keyword “COVID-19” in both Japanese and English were extracted. In addition to the time series analysis and LDA topic modeling previously described the correlation between all HPV vaccine-related tweets and the tweets incorporating the “COVID-19” keywords was examined. Moreover, specific time points where the total number of tweets and the tweets containing “COVID-19” keywords exhibited simultaneous increases were investigated. To estimate the percentage of tweets pertaining to the pivotal events, 100 tweets were randomly selected thrice from the time points. The number of tweets related to the paramount event was verified by three different volunteers, and the percentage of related tweets was calculated by averaging the three results and computing the confidence interval (CI). To gain a deeper understanding of the spikes in the data, the influence of COVID-19 vaccine-related pivotal events on the peak time points was also assessed.

Furthermore, we investigated the potential causality between the stance towards the HPV vaccine and the stance towards the COVID-19. Specifically, we analyzed tweets containing “COVID-19” keywords and expressing either an advocate or opposed stance. We employed a logic analysis by LLM to determine whether the tweet utilized the HPV vaccine as an illustrative example to convey the author’s perspective on the COVID-19, vice versa, or neither. The Claude-3-opus model was directly employed for inference, with the prompt instructing it to read the tweet and provide an output of “HPV to COVID” if the tweet used the HPV vaccine as an example to express an idea about the COVID-19 vaccine, “COVID to HPV” if the reverse was true, or “Not related” if neither was applicable. The classified tweets were tallied weekly and visually represented to demonstrate the results.

## Results

### Model Selection

The comparison of all models we tried are shown in Figure 2. F1 scores of finetuned larger models get better performance in most cases, except for Llama 3.1. This may because it was not instruction tuned on Japanese corpora in advance. Finetuned Gemini 1.0 pro got the best results in our experiment, and we decided to do the following steps based on Gemini 1.0 pro. Detailed results are in Appendix 2 and 3.

**Figure 2.**
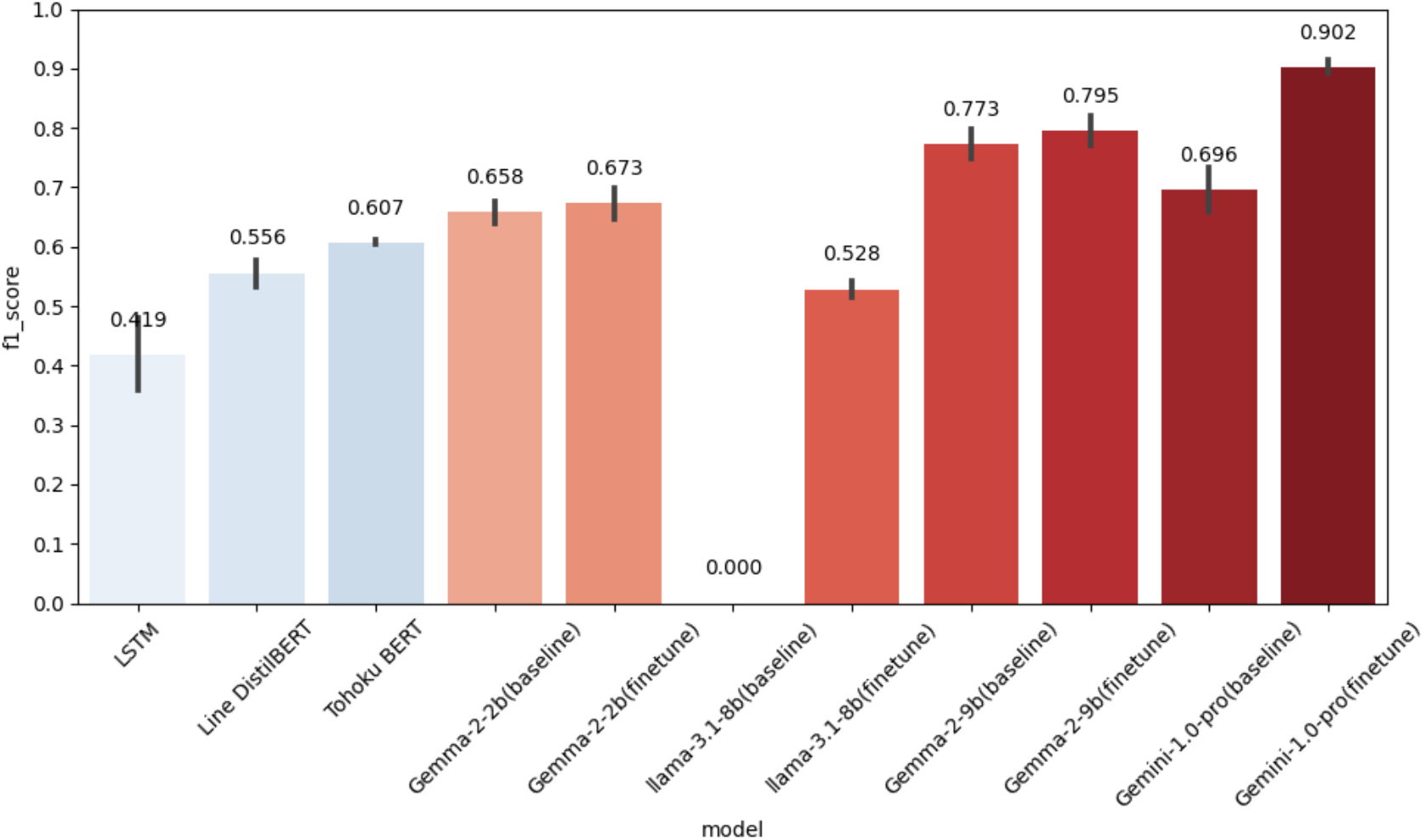
Comparison of weighted F1 scores among all the models on 300 test data. Bars show the average and standard deviation on three test datasets selected by different random seeds. The Numbers above the bars show the average of weighted F1 scores.

As the allowed number of tweets for finetuning Gemini 1.0 pro is fixed (<500), we experimented with different data ratios to find the most effective category ratio to finetune the model. The results indicated that the model performed best when the ratio of advocate, opposite, and unknown categories was 150:200:150. This ratio consistently appeared in validation experiments, achieving the highest average F1-score of 0.968, as shown in Figure 3. Detailed model performance for each ratio can be found in Appendix 2.

**Figure 3.**
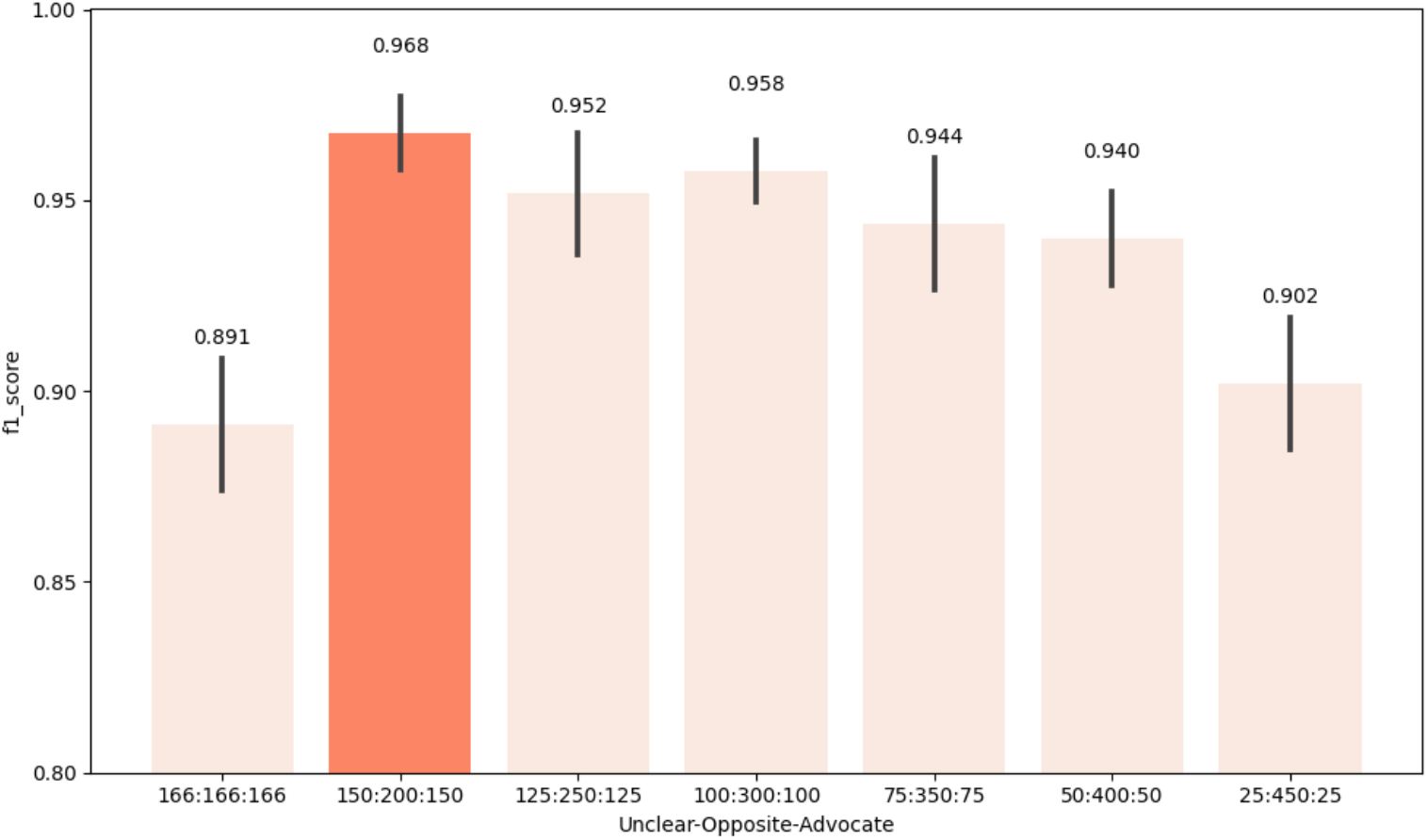
Comparison of weighted F1 scores among models trained with different ratios of categories in the training data. Bars show the average and standard deviation on three test datasets selected by different random seeds. The Numbers above the bars show the average of weighted F1 scores.

After conducting five rounds of random data selection and fine-tuning using the optimal data ratio, the performance of the fine-tuned Gemini 1.0 Pro model was evaluated on the remaining labeled data. The evaluation results showed a significant improvement in precision, recall, and F1-score for the fine-tuned model. The average F1-score of the three best-performing models was 0.924, with a precision of 0.928 and recall of 0.924. In comparison, the baseline performance of the untuned Gemini 1.0 Pro model on the same test dataset showed an F1-score of 0.781, precision of 0.829, and recall of 0.796. This comparison clearly demonstrates the importance of fine-tuning in enhancing the model’s performance for specific tasks.

When classifying the unlabeled Japanese tweets, the results from the three best models were mostly consistent. Among all tweets, 86.85% received three identical result labels, and 13.06% received two identical result labels. In cases where all three labels were completely inconsistent, these tweets were classified as unknown, accounting for 0.09% of all tweets.

### Time series analysis of stance

We first analyzed the monthly trend of public stances towards HPV vaccine from 2011 to 2021, as shown in Figure 4. Tweets for advocate and opposed stances both increased rapidly within a decade, while advocation gradually dominated the stances.

**Figure 4.**
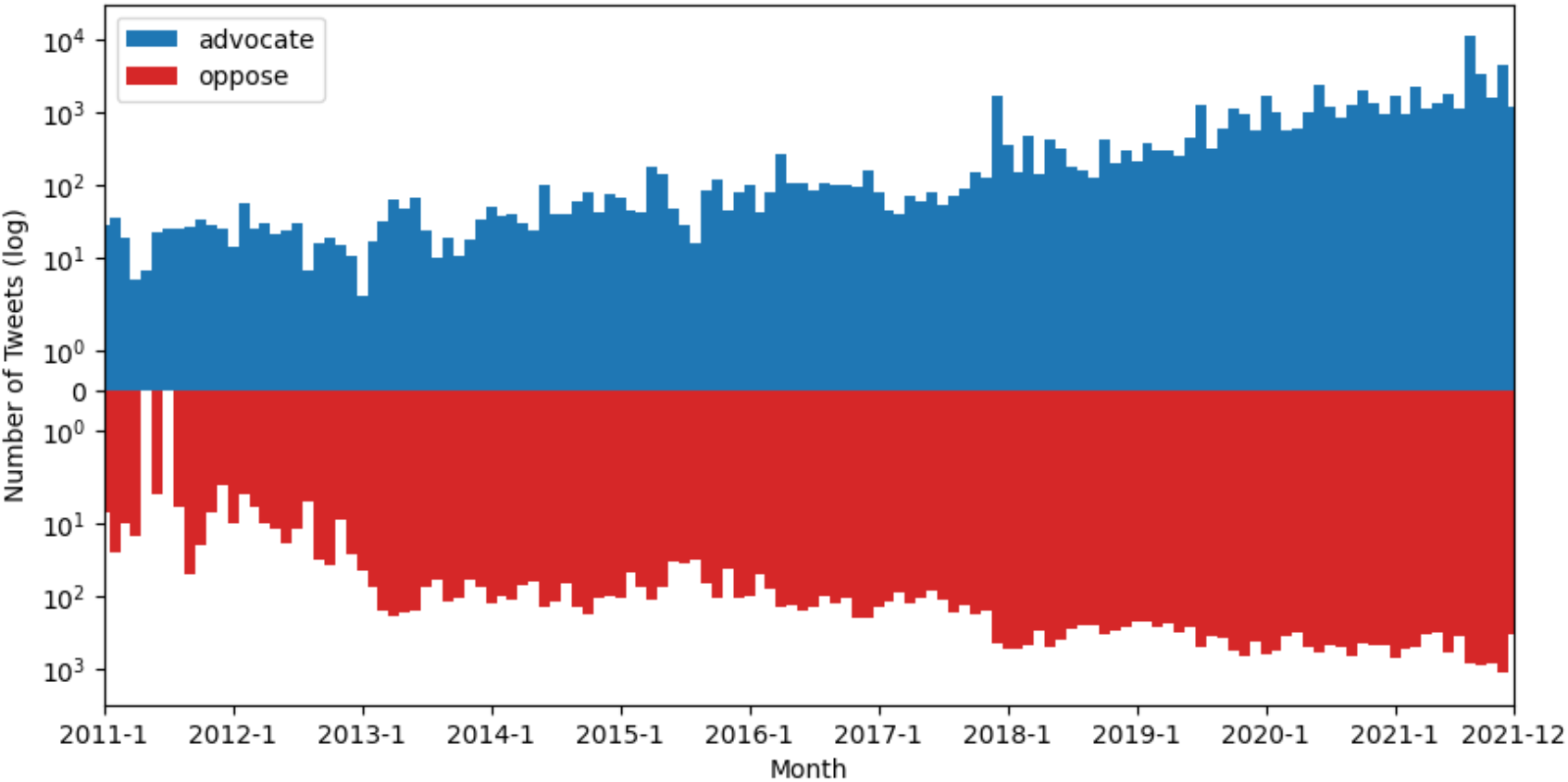
Log-scaled monthly number of tweets of advocate and oppose stances.

The time series analysis of difference between advocate and opposed stances is visualized in Figure 5, unveiled several pivotal junctures marked by substantial shifts in the disparity between pro- and anti-HPV vaccination stances, suggesting notable fluctuations in public opinion. Overall, we can see there are three periods of fluctuations and three periods being relatively stable in stances. Employing the PELT algorithm, we successfully discerned structural change points within the data, aligning closely with significant shifts in public attitudes towards HPV vaccines. Notably, these change points were observed in 2013, 2016, and 2020, coinciding with key public health events and policy developments. In 2013, the Japanese government’s decision to suspend its recommendation for the HPV vaccine [54] precipitated a marked decline in public confidence. Subsequently, in late 2016, widespread discourse surrounding vaccine safety was triggered by legal action taken by individuals alleging adverse side effects [55]. Finally, 2020 witnessed a resurgence in public advocacy for HPV vaccination, with numerous petitions urging the government to reinstate active recommendations, prompting a policy review [56].

**Figure 5.**
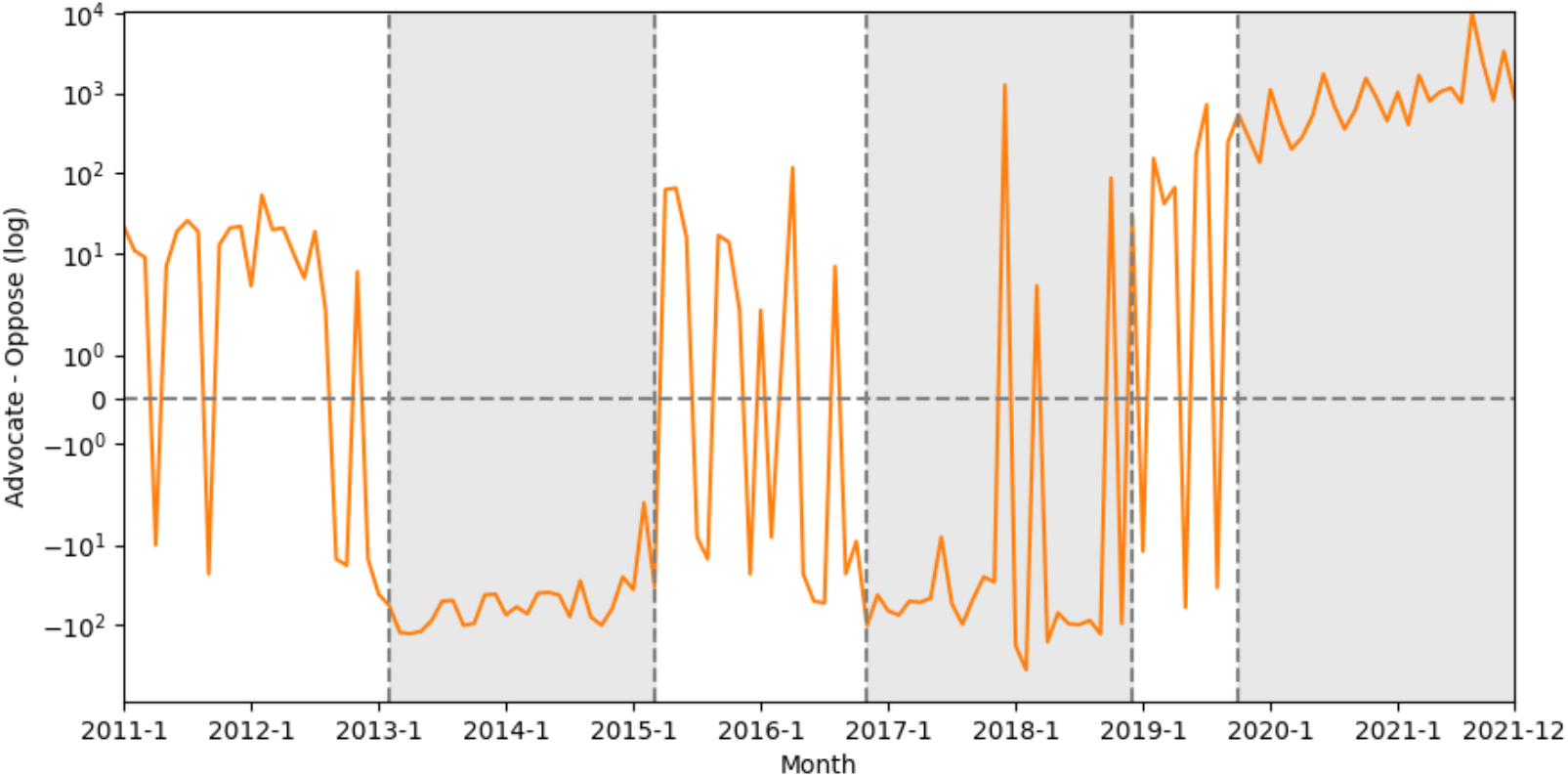
Monthly differences between number of tweets of advocate and oppose stances. The significant low and high periods based on change points detected by PELT are marked in gray.

### LDA topic modeling analysis

We applied LDA modeling on the tweets of advocate, oppose, unknown stances separately. The best topic number for advocate and unknown are three, and that for oppose is four. The details of top words and weights are shown in Appendix 4. Then we calculated the ratio of the monthly expectations of tweets belonging to different topics to get fine-grained insight to the weight of topics in each stance.

The distribution of various themes within the “advocate” stance is illustrated in Figure 6. The proportion of “Scientific and media discourse on HPV Vaccine Safety” (Topic 1) underwent a significant increase until 2015, subsequently experiencing a gradual decline. Conversely, the proportion of “HPV vaccine effectiveness and broader public health measures” (Topic 2) diminished, reaching its lowest point around 2015, after which it exhibited a consistent increase. The proportion of “Policy and advocacy for HPV vaccination promotion” (Topic 3) remained relatively stable, although a slight upward trend has been observed since 2019.

**Figure 6.**
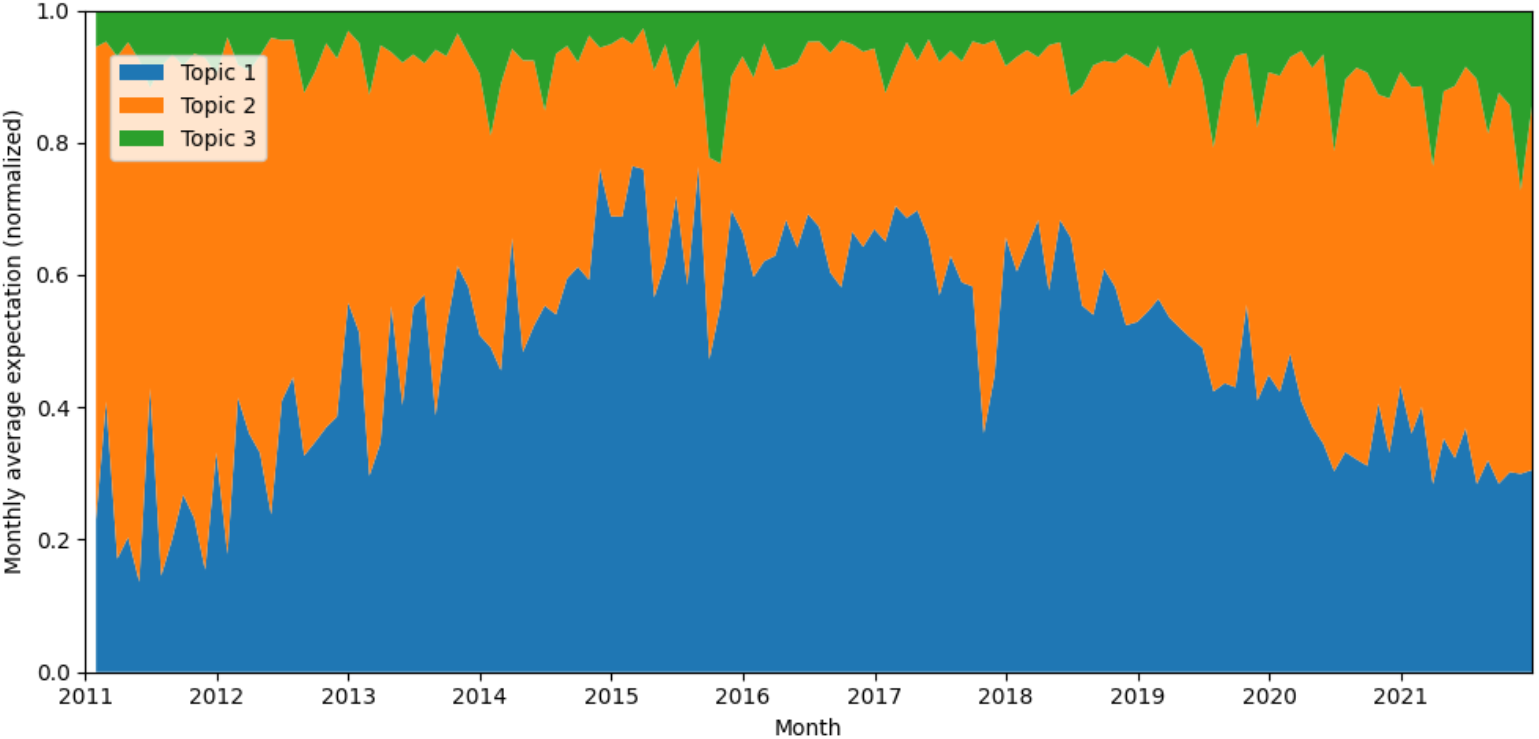
Monthly change of ratio of different topics in tweets of advocate stance from 2011 to 2021.

In the analysis of tweets expressing the “opposing” stance in Figure 7, “Skepticism and opposition to vaccination” (Topic 2) demonstrated notable peaks in weight during the years 2013 and 2015, followed by a gradual decline and eventual stabilization of topic weight.

**Figure 7.**
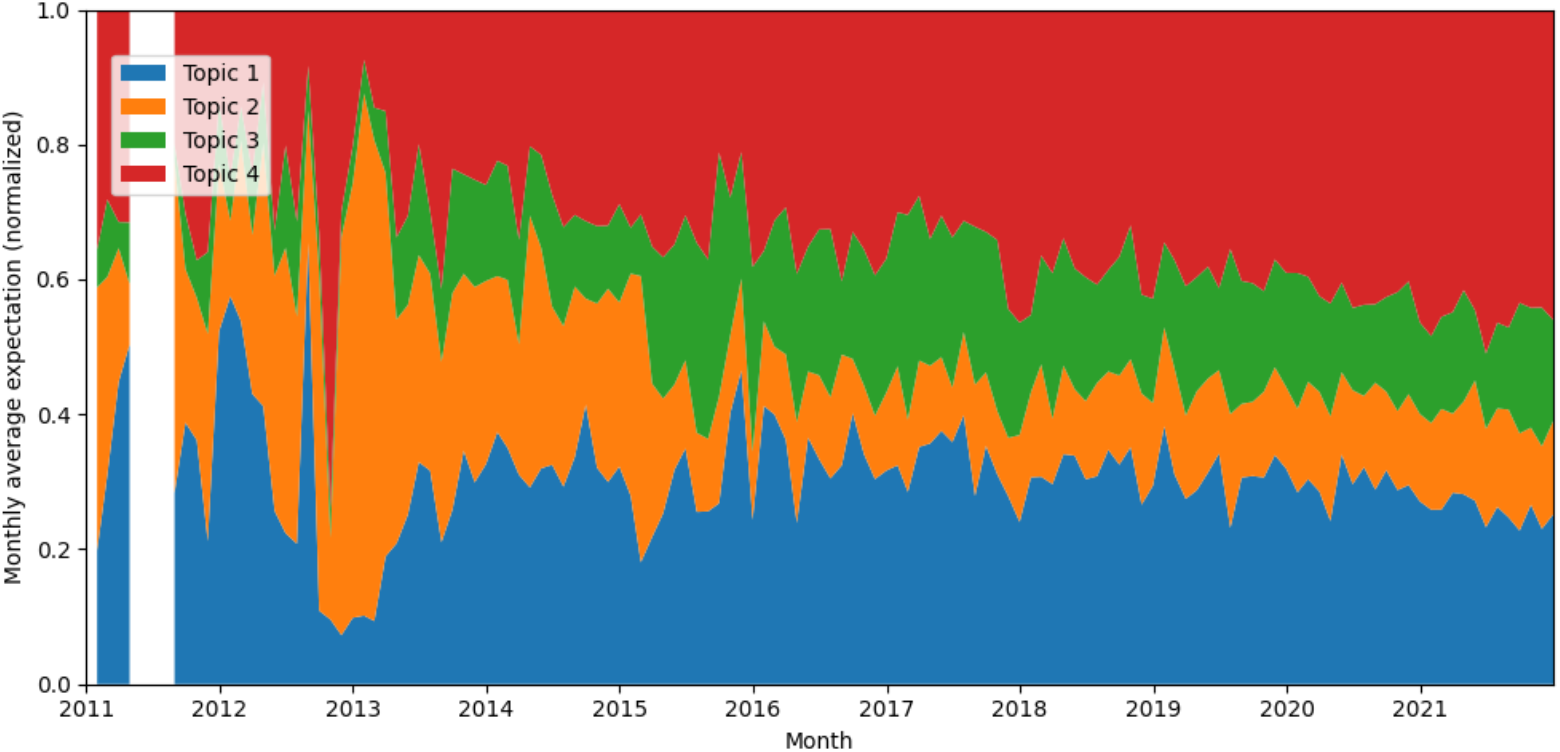
Monthly change of ratio of different topics in tweets of opposed stance from 2011 to 2021.

The proportion of “Scientific warnings and public health risks” (Topic 4) experienced a surge in late 2012, followed by a sustained increase throughout the decade, ultimately becoming the dominant theme within the opposing stance.

In the tweets classified as “unknown” by stance shown in Figure 8, “HPV Vaccine Efficacy and Public Health Initiatives” (Topic 2) was the dominant theme prior to the year 2013, then declined thereafter. Conversely, the proportion of tweets expressing “Opposition, Misinformation, and Activism Surrounding HPV Vaccination” (Topic 3) displayed a gradual increase and has become the predominant theme since 2018. Notably, the theme of “HPV Vaccine Safety and Governmental Oversight” (Topic 1) exhibited a gradual increase throughout the entire period under consideration.

**Figure 8.**
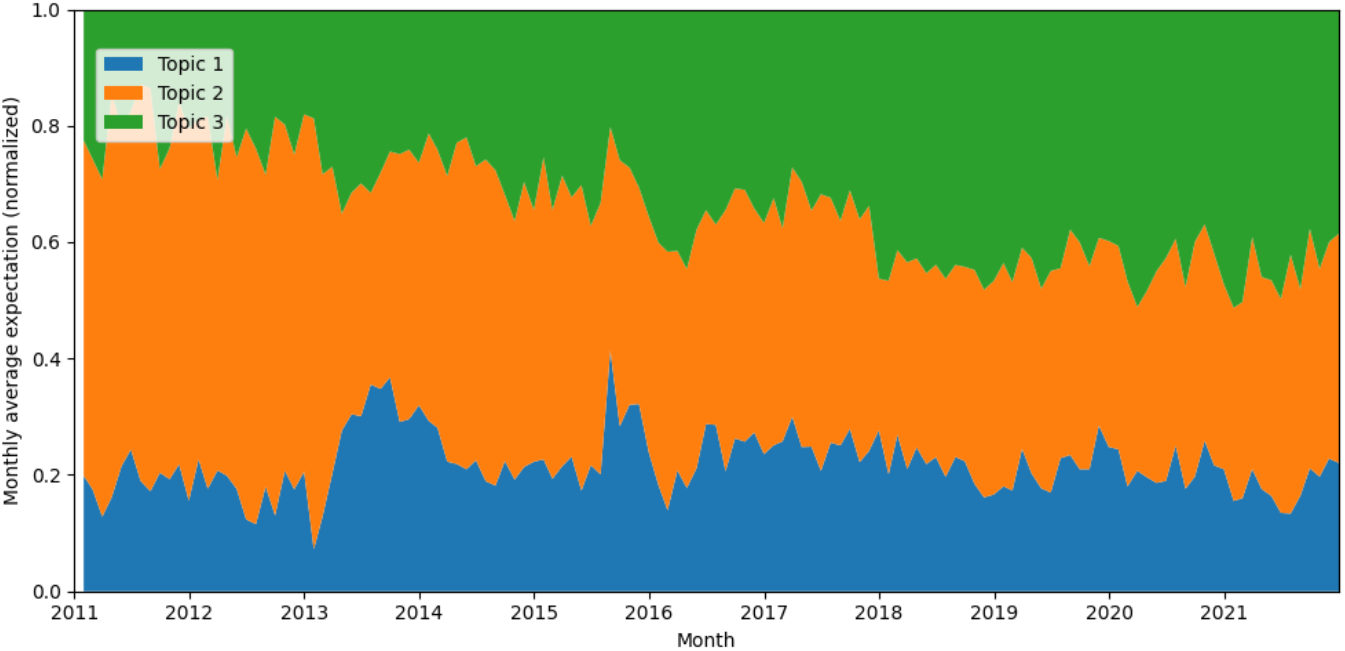
Monthly change of ratio of different topics in tweets where the stances are unknown from 2011 to 2021.

Then we applied misinformation detection on some topics. In tweets of opposed stance, we identified “Scientific warnings and public health risks” (Topic 4) as the most likely to contain misinformation about the HPV vaccine. We extracted tweets with over 80% probability of belonging to this topic, yielding a total of 3,001 tweets. The misinformation classification results, obtained using the Claude-3-opus, are presented in Figure 9. The classification accuracy, determined through random sampling and verification conducted thrice, ranged from 89.74% to 100% (CI=95%). Our findings reveal that the prevalence of misinformation increased in most years, with exceptions in 2014 and 2015. The ratio of misinformation in HPV vaccine-related tweets initiated with a sharp spike in 2012, followed by a rapid decline to its nadir in 2015. Subsequently, the ratio gradually increased, reaching a secondary but albeit lower peak in 2018, after which it began a gradual decline.

**Figure 9.**
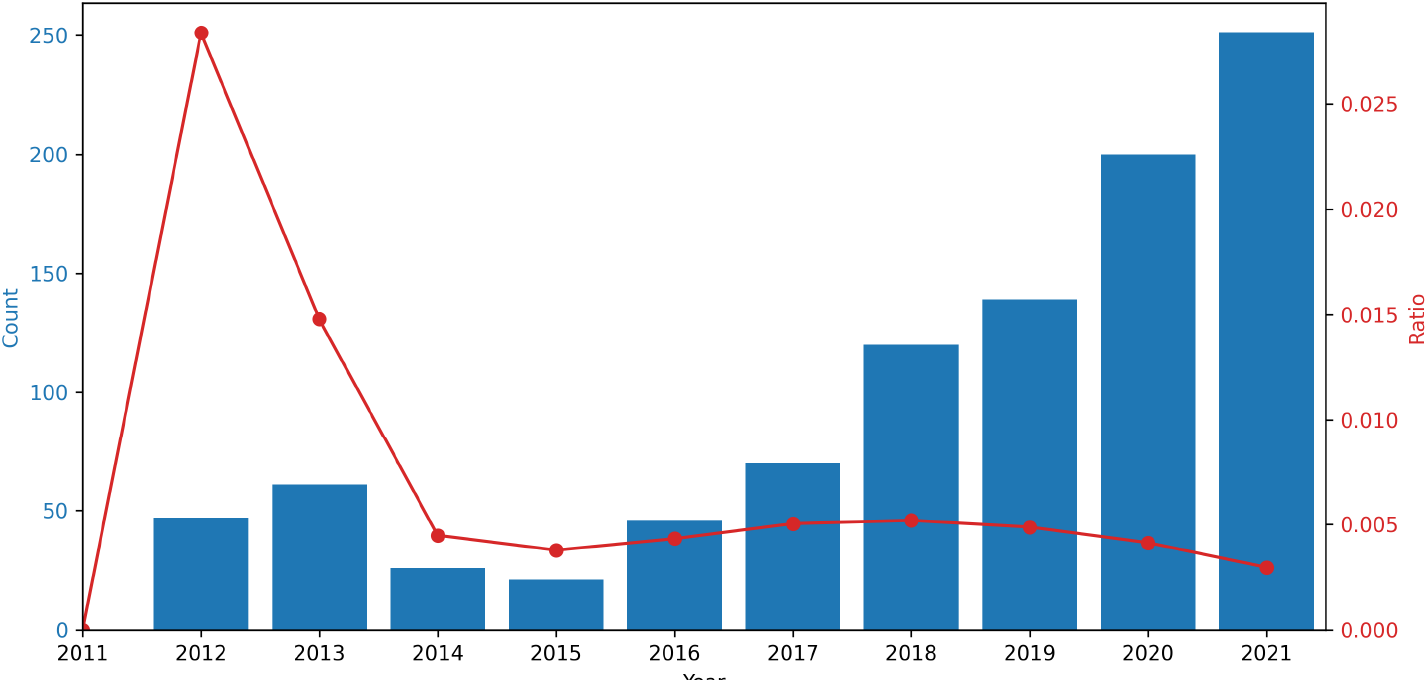
Count and ratio of tweets classified as misinformation. The bars shows the total number of tweets found as misinformation each year, and the line shows the ratio of the tweets containing misinformation within the total number of tweets each year.

### Relationship between HPV vaccination and COVID-19

We conducted a weekly time series analysis on the stances of tweets containing the COVID-19 keywords, as illustrated in Figure 10. There are no tweets with “COVID-19” keyword in the first two weeks of 2020, so all the calculations began from the 3rd week. The Pearson correlation coefficient (*r*) was 0.793 (p = 1.59*10^−23^), indicating a relatively high correlation between all tweets and those containing “COVID-19” keywords. The time lag for the highest cross correlation was zero. Several peaks in both all tweets and tweets containing “COVID-19” keywords were observed in 2021 (week 3, week 34∼35, and week 45), with the 34∼35th week exhibiting the highest peak during the entire study period, which coincided with a campaign to call for resumption of active HPV vaccination recommendations [57] (37.0%∼39.8% HPV vaccine-related tweets; 1.5% ∼ 9.1% tweets with “COVID-19” keywords; CI=95%).

**Figure 10.**
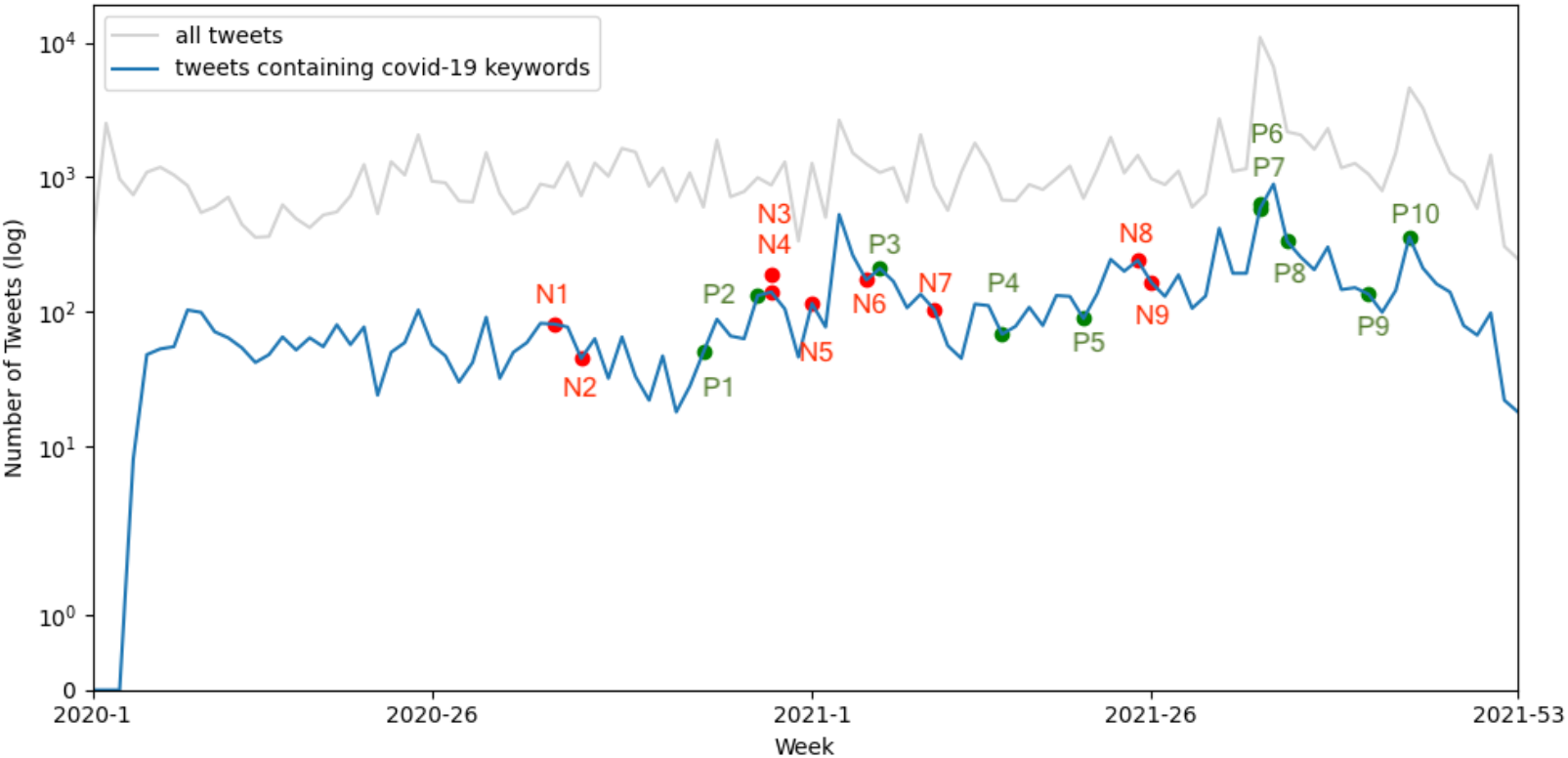
Number of all tweets related to HPV vaccine (gray), and the tweets containing the “COVID-19” keyword (blue), with key events related to COVID-19 vaccine marked. The points beginning with “N” are negative key events that may lead to opposition to COVID-19 vaccines, and the points beginning with “P” are positive key events that may lead to advocacy to COVID-19 vaccines.

A comparative analysis was performed to examine the differences between advocate and opposed stances in both the overall tweet corpus and the subset containing the “COVID-19” keyword, as illustrated in Figure 11. The *r* between advocate and oppose sentiment differences is substantial (r= 0.723, p = 6.09*10^−18^), with no temporal lag observed in the highest cross-correlation. In the broader context of all HPV vaccine-related tweets, advocate sentiment generally prevailed over oppose sentiment. However, for tweets containing “COVID-19” keywords, the dominant stance fluctuated over time. Both datasets exhibited a pronounced peak in advocacy during weeks 34-35 of 2021, while oppose sentiment dominated in week 22 of 2020. This pattern was consistent across the entire tweet corpus and the “COVID-19” keyword subset.

**Figure 11.**
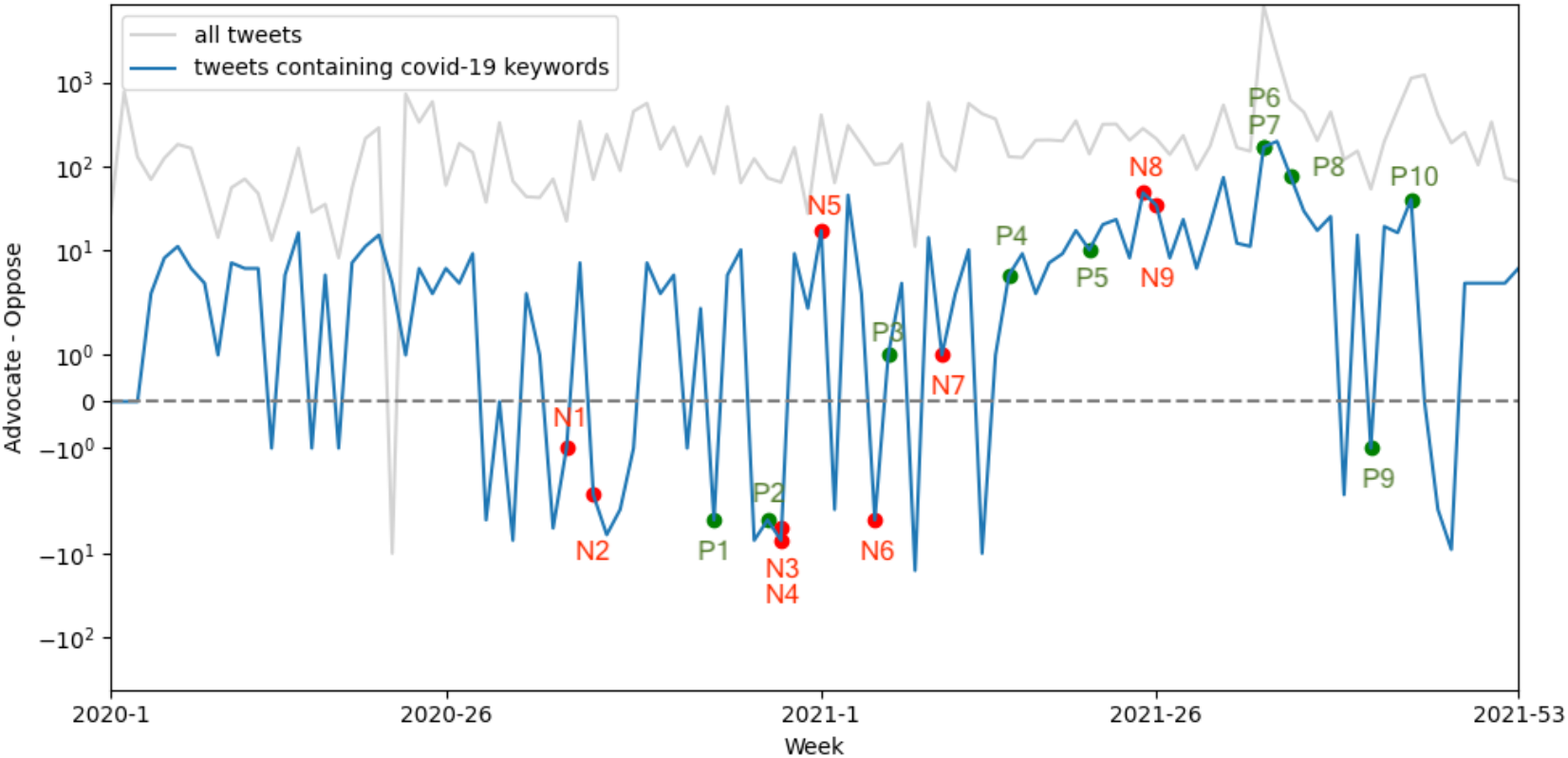
The difference between tweets of advocate stance and oppose stance for all HPV vaccine tweets (gray) and tweets containing the “COVID-19” keyword (blue), with key events related to COVID-19 vaccine marked. The points beginning with “N” are negative key events that may lead to opposition to COVID-19 vaccines, and the points beginning with “P” are positive key events that may lead to advocacy to COVID-19 vaccines.

We additionally assessed the impact of COVID-19 vaccine-related events on the volume of all HPV vaccine-related tweets and tweets containing the keywords “COVID-19.” The details of the key events were gathered from [58] and are provided in Table 1 of Appendix 4. As illustrated in Figure 10, it is evident that the key events do not consistently result in peaks in tweet volume for both all HPV vaccine-related tweets and tweets containing the keywords “COVID-19.” Furthermore, as depicted in Figure 11, the positive or negative events do not always lead to an increase in the number of supportive or opposing stances.

LDA was conducted on tweets containing COVID-19 keywords of advocating and opposing stances separately. The detailed results are presented in Appendix 4. To investigate nuanced changes in topic weights over time, the expectation of tweets belonging to different topics for both advocate and oppose stances was calculated and visualized in Appendix 4. Analysis of this data revealed no significant changes in the proportion of different topics during 2020 and 2021.

Finally, we investigated the potential causal relationship between attitudes towards HPV and COVID-19 vaccines by logic analysis. To evaluate the accuracy of Claude-3-opus doing the classification task without finetuning, we randomly sampled 100 tweets three times and let three volunteers judge the correctness. The accuracy is 72.92%∼91.74%, CI=95%. The weekly classification results are illustrated in Figure 12. The “HPV to COVID” category predominated throughout the entire study period for both supportive and opposing stances. Notably, tweets advocating for vaccines consistently outnumbered those opposing them across all categories.

**Figure 12.**
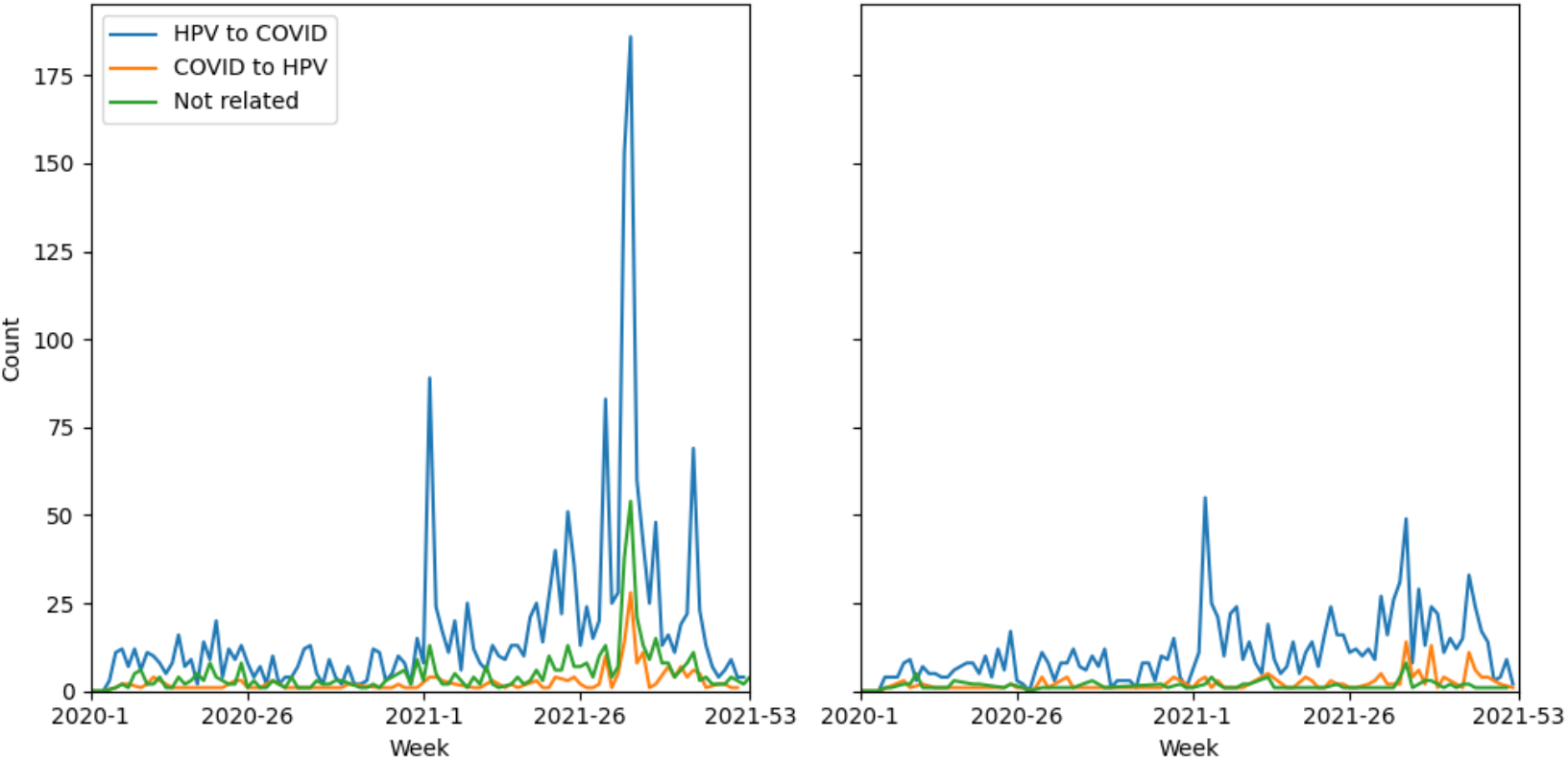
Weekly number of tweets with “COVID-19” keywords, using HPV vaccine as example to share stance to COVID-19 vaccine (“HPV to COVID”), using COVID-19 vaccine as example to share stance to HPV vaccine (“COVID to HPV”), or neither (“Not related”). Left: result on tweets with advocate stance; Right: result on tweets with opposed stance.

## Discussion

### Principal Results

In this work, we employed traditional NLP models and LLMs to perform stance analysis on social media content, focusing on the public discourse around the HPV vaccine. Our results demonstrate that stance analysis using LLMs provides a more nuanced understanding compared to traditional sentiment analysis. Unlike traditional sentiment analysis, which captures general emotional tones, LLMs can identify specific stances—such as supportive, opposing, or neutral positions—allowing for a deeper comprehension of public attitudes and decision-making processes. Moreover, the successful application of LLMs highlights their potential as powerful tools for social media analysis, providing deep insights into public health discourse by capturing more complex logic like causality and context-specific nuances. These results not only validate the effectiveness of our methodology but also provide strong empirical support for using large language models in similar tasks in the future.

In the context of our findings about HPV vaccine, we observed increase in public advocacy and vaccine confidence towards HPV vaccine, although fair propagation and communication is still a key factor to further drive the HPV vaccination. The WHO’s 3Cs model (Confidence, Complacency, Convenience) offers a comprehensive framework for understanding the factors influencing vaccine hesitancy in Japan.

Confidence pertains to trust in the vaccine’s safety, efficacy, and the healthcare system responsible for its administration, and plays the key role in the stance towards HPV vaccine in Japan. In our analysis, we observed significant fluctuations in public stances towards the HPV vaccine, which were closely linked to government policy decisions and media coverage. The 2013 suspension of HPV vaccine recommendations by the Japanese government was a key moment that led to a sharp decline in public trust. This decline can be attributed to increased public uncertainty and fears regarding vaccine safety, as the time series analysis shows. The LDA topic modeling results also support this, showing a decline in the proportion of tweets related to “HPV Vaccine Efficacy and Public Health Initiatives” (Topic 2) where the stance is unknown after 2013, while “Opposition, Misinformation, and Activism Surrounding HPV Vaccination” (Topic 3) gradually became the predominant theme since 2018, highlighting growing public distrust and misinformation. Conversely, the resurgence in advocacy in 2020 reflected an improvement in public confidence, likely influenced efforts to reinstate vaccine recommendations. This is reflected in the increased proportion of tweets related to “Scientific and Media Discourse on HPV Vaccine Safety” within the ‘advocate’ stance, which showed a significant increase until 2015, followed by a gradual decline, indicating ongoing advocacy efforts. Our findings illustrate that confidence is highly sensitive to policy decisions and that restoring public trust after a decline requires consistent and clear communication efforts.

Furthermore, we explored the role of misinformation in undermining public confidence in the HPV vaccine. The spike in the ratio of tweets containing misinformation in 2012 likely contributed to the significant drop in vaccine confidence observed in the following year, which aligns with findings from other studies [22,31,59]. From 2014 onwards, the ratio of misinformation exhibited slight changes but remained consistently present, and the overall number of misinformation tweets increased in most years, indicating the persistent and long-lasting impact of misinformation. Addressing misinformation is therefore essential for restoring and maintaining public trust in vaccines, as unchecked misinformation can severely erode public confidence and contribute to vaccine hesitancy.

Complacency refers to the perceived necessity of vaccination, which diminishes when individuals consider the risk of vaccine-preventable diseases to be low. Eric et al. found that “not necessary” was consistently among the top reasons cited by parents for not vaccinating their children against HPV between 2010 and 2020 [60]. The topic “HPV Vaccine Efficacy and Public Health Initiatives” under the ‘unknown’ stance experienced a sharp decline post-2013, indicating diminished public engagement and perceived necessity. The perceived low risk of HPV, combined with concerns about side effects, may contributed to a reduced sense of urgency regarding vaccination. However, our time series analysis showed that complacency began to decrease in 2020 as advocacy efforts intensified, and the public became more aware of the risks of HPV and the benefits of vaccination. The upward trend in the topic “HPV Vaccine Effectiveness and Broader Public Health Measures” within the ‘advocate’ stance since 2015 further reflects this shift towards increased awareness and decreasing complacency. This shift underscores the importance of maintaining proactive communication about the risks of vaccine-preventable diseases to counteract complacency.

Convenience encompasses factors such as the availability, affordability, and accessibility of vaccination. In our study, the importance of convenience became evident when analyzing periods of increased advocacy for HPV vaccination. For example, the resurgence of advocacy in 2020 coincided with governmental initiatives aimed at improving access to the HPV vaccine, including the reintroduction of public recommendations and the enhancement of vaccination services. This alignment is reflected in the relatively stable proportion of tweets related to ‘Policy and Advocacy for HPV Vaccination Promotion’ within the ‘advocate’ stance, which exhibited a slight upward trend since 2019. These changes may encourage efforts to facilitate access to vaccination services, and play a critical role in influencing public willingness to get vaccinated over time.

Beside the findings about HPV vaccine, we also noticed that the stances towards the HPV vaccine may influence stances towards the COVID-19 vaccine during the COVID-19 pandemic. Specifically, our logic analysis revealed that increased confidence in the HPV vaccine may lead to higher confidence in the COVID-19 vaccine. Tweets categorized under ‘HPV to COVID’—which used the HPV vaccine as a reference point for the COVID-19 vaccine stance—predominated throughout the study period for both supportive and opposing stances. Notably, advocacy for vaccines consistently outnumbered opposition across all categories, suggesting that rising confidence in one vaccine may positively impact confidence in other vaccines.

For future work, we plan to explore why LLMs outperform traditional models, focusing on factors like model size and external information. We are also considering building a LLM to identify misinformation about HPV vaccine with higher accuracy. These efforts will refine our understanding of LLMs and inform better public health strategies.

### Limitations

We admit that our work has many limitations, including the fact that social media users may not represent the entire population, which limits the generalizability of our findings. Additionally, the models may introduce classification errors, particularly when dealing with nuanced or ambiguous stances and logics. Furthermore, the keywords used for collecting data may have led to the omission of relevant tweets or added irrelevant tweets, potentially impacting the comprehensiveness of our analysis.

### Comparison with Prior Work

Our work stands out from prior studies due to the scale and depth of the dataset, which includes ten years of tweets in Japanese, providing a comprehensive view of public attitudes over time. We applied both traditional NLP methods and cutting-edge LLMs, allowing us to analyze nuanced shifts in public sentiment and stance with greater precision. Unlike traditional surveys, which provide snapshots of attitudes, our approach captures real-time changes in public discourse. Additionally, compared to studies relying on Google search trends, our analysis offers more fine-grained insights into specific stances and topics, highlighting not just information-seeking behavior, but the underlying attitudes that drive vaccine hesitancy or advocacy.

## Conclusions

This study highlights the effectiveness of LLMs for stance analysis in understanding public attitudes towards HPV vaccination. By applying the WHO’s 3Cs model, we contextualized the complex factors influencing stances towards HPV vaccine. Public confidence fluctuated significantly in response to government actions and media coverage, showing the sensitivity of trust to policy decisions, while complacency was impacted by perceived risks and proactive advocacy. Convenience was crucial for improving vaccine accessibility, shaping public willingness to get vaccinated. Moreover, our findings suggest that confidence in one vaccine, such as HPV, may influence confidence in others, like COVID-19, highlighting interconnected public health narratives. Addressing misinformation, enhancing communication, and improving accessibility remain key strategies for building trust and reducing hesitancy. Future public health strategies can benefit from these insights to design effective interventions aimed at boosting vaccine confidence and uptake.

## Data Availability

All data produced in the present study are available upon reasonable request to the authors

## Acknowledgements

This work was supported by the JST SPRING (grant number JPMJSP2110) and Google PhD fellowship.

## Abbreviations

BERT: Bidirectional Encoder Representations from Transformers
CI: confidence interval
DL: deep learning
HPV: human papillomavirus
LDA: latent Dirichlet allocation
LLM: large language model
LSTM: long short-term memory
MHLW: Japanese Ministry of Health, Labour and Welfare
NLP: natural language processing
PELT: pruned exact linear time
QLoRA: Quantization and Low-Rank Adaptation

